# Exaggerated Right Ventricular Dysfunction in Females with Group 3 Pulmonary Hypertension

**DOI:** 10.1101/2021.04.16.21255516

**Authors:** Sasha Z. Prisco, Felipe Kazmirczak, Kurt W. Prins, Thenappan Thenappan

**Author notes:** **Authors for Correspondences:** Thenappan Thenappan, MD, Associate Professor of Medicine, Cardiovascular Division, University of Minnesota, Twitter: @thenappanMD, Kurt Prins, MD, PhD, Assistant Professor of Medicine, Cardiovascular Division, Lillehei Heart Institute, University of Minnesota, Twitter: @kurt_prins.

## Abstract

Group 3 pulmonary hypertension (PH) patients have disproportionate right ventricular dysfunction (RVD) as compared to pulmonary arterial hypertension (PAH) patients, but the cause of the divergent RV phenotypes is unknown. One potential mechanism may be biological sex as females have better RV function than males. However, the combined effects of PH type and sex on RV function are unexplored. Therefore, we evaluated how sex and PH etiology modulated RVD in a single-center cohort study. Male sex was not associated with significant differences in RV function when comparing PH etiologies. However, female Group 3 patients had more pronounced RVD than female PAH patients. In particular, Group 3 females had marked reduction in RV function when pulmonary vascular resistance was matched. Group 3 females were older than PAH females, but the exaggerated RVD was still observed in postmenopausal (age≥55) Group 3 females. This suggests lung disease exacerbates RVD in Group 3 females.

---

Pulmonary hypertension (PH) due to chronic lung disease (Group 3 PH) is the second most common cause of PH(1), and Group 3 PH patients have the worst survival rates of all the PH etiologies(2). An important prognostic factor in Group 3 PH is the presence of right ventricular dysfunction (RVD), as RVD results in heightened rates of heart failure hospitalization and death(3). Interestingly, Group 3 PH patients have worse RV function despite having less severe PH when compared to pulmonary arterial hypertension (PAH) patients(4). Currently, the mechanisms underlying the disproportionate RVD in Group 3 PH patients are unknown. A deeper understanding of this observation is important because this patient population has limited treatment options.

One potential explanation for the disproportionate RVD in Group 3 PH may be biological sex as Group 3 PH does not have a female predominance like PAH does(5), and multiple studies reveal females have superior RV function when compared to males(6). However, the potential synergetic effects of PH etiology and biological sex on RV function in PH patients are unexplored. Therefore, we examined how PH type impacted RV function in males and females.

## Methods

Group 1 PAH and Group 3 PH patients were identified from the Minnesota PH Repository(7) and were defined according to the World Health Organization criteria as outlined by the 6th World Symposium on PH(8). We used pulmonary function tests, high resolution computed tomography, and sleep studies to define Group 3 PH. RV fractional area change (RVFAC) was quantified from offline transthoracic echocardiograms by two reviewers (FK and KWP). Invasive hemodynamics confirmed PH diagnosis (mean pulmonary arterial pressure [mPAP]>20 mm Hg). We evaluated the relationship between RVFAC and pulmonary vascular resistance (PVR) based on sex and PH etiology. PVR was defined as (mPAP - pulmonary capillary wedge pressure)/ cardiac output. Pulmonary arterial compliance (PAC) was calculated as stroke volume divided by pulmonary arterial pulse pressure. Statistical analysis was performed on GraphPad Prism version 9.0.0. Unpaired *t*-test compared means of two groups. If variance was unequal, Mann-Whitney U test was completed. Fisher’s exact test evaluated differences in categorical variables. Data are presented as mean±standard deviation.

## Results

We identified 217 PAH (60 males and 157 females) and 233 Group 3 PH patients (105 males and 128 females) for our analysis (**Supplemental Table**). Group 3 PH patients were older than PAH patients in both male (Group 3: 67±11 years vs. Group 1: 53±13 years, *p*<0.0001) and female sexes (Group 3: 64±11 vs. Group 1: 54±18 years, *p*<0.0001) (**Table**). Male Group 3 PH patients had lower mPAP (Group 3: 40±10 vs. Group 1: 47±16 mmHg, *p*=0.007) with a trend for lower PVR (Group 3: 6.5±3.4 vs. Group 1: 7.5±4.2 WU, *p*=0.09) compared to PAH males (**Table**). Although there was no difference in mPAP between female Group 3 and PAH patients, female Group 3 PH patients had lower PVR (Group 3: 7.2±3.8 vs. Group 1: 9.5±5.8 WU, *p*=0.002). There were no differences in PAC between Group 1 and 3 PH patients within the same sex. In summary, Group 3 patients had either similar or less severe pulmonary vascular disease compared to PAH patients of the same sex.

**Table:**
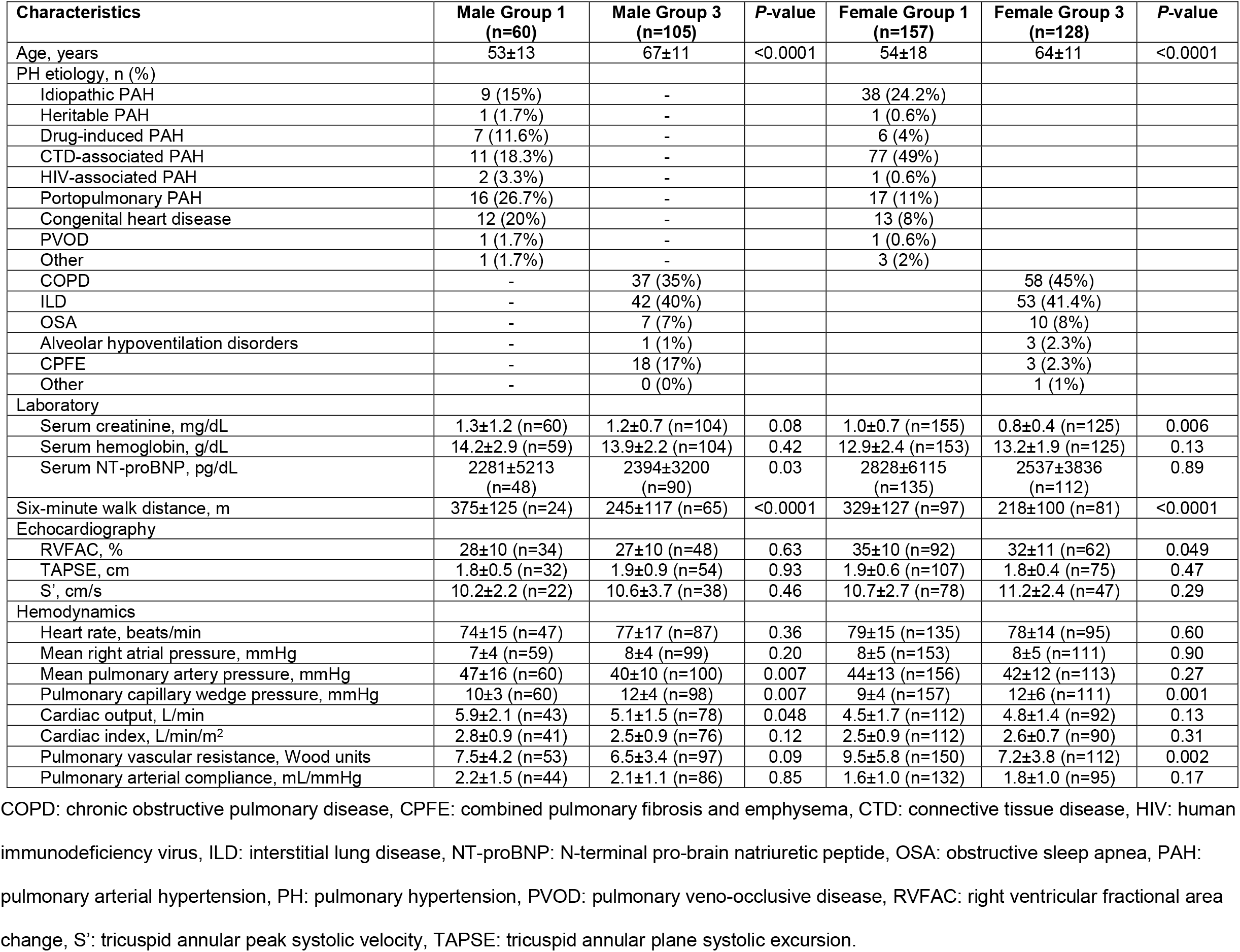
Clinical characteristics of the Group 1 PAH and Group 3 PH cohorts by sex.

| Characteristics | Male Group 1<br>(n=60) | Male Group 3<br>(n=105) | P-value | Female Group 1<br>(n=157) | Female Group 3<br>(n=128) | P-value |
| --- | --- | --- | --- | --- | --- | --- |
| Age, years | 53±13 | 67±11 | <0.0001 | 54±18 | 64±11 | <0.0001 |
| PH etiology, n (%) |  |  |  |  |  |  |
| Idiopathic PAH | 9 (15%) | - |  | 38 (24.2%) |  |  |
| Heritable PAH | 1 (1.7%) | - |  | 1 (0.6%) |  |  |
| Drug-induced PAH | 7 (11.6%) | - |  | 6 (4%) |  |  |
| CTD-associated PAH | 11 (18.3%) | - |  | 77 (49%) |  |  |
| HIV-associated PAH | 2 (3.3%) | - |  | 1 (0.6%) |  |  |
| Portopulmonary PAH | 16 (26.7%) | - |  | 17 (11%) |  |  |
| Congenital heart disease | 12 (20%) | - |  | 13 (8%) |  |  |
| PVOD | 1 (1.7%) | - |  | 1 (0.6%) |  |  |
| Other | 1 (1.7%) | - |  | 3 (2%) |  |  |
| COPD | - | 37 (35%) |  |  | 58 (45%) |  |
| ILD | - | 42 (40%) |  |  | 53 (41.4%) |  |
| OSA | - | 7 (7%) |  |  | 10 (8%) |  |
| Alveolar hypoventilation disorders | - | 1 (1%) |  |  | 3 (2.3%) |  |
| CPFE | - | 18 (17%) |  |  | 3 (2.3%) |  |
| Other | - | 0 (0%) |  |  | 1 (1%) |  |
| Laboratory |  |  |  |  |  |  |
| Serum creatinine, mg/dL | 1.3±1.2 (n=60) | 1.2±0.7 (n=104) | 0.08 | 1.0±0.7 (n=155) | 0.8±0.4 (n=125) | 0.006 |
| Serum hemoglobin, g/dL | 14.2±2.9 (n=59) | 13.9±2.2 (n=104) | 0.42 | 12.9±2.4 (n=153) | 13.2±1.9 (n=125) | 0.13 |
| Serum NT-proBNP, pg/dL | 2281±5213<br>(n=48) | 2394±3200<br>(n=90) | 0.03 | 2828±6115<br>(n=135) | 2537±3836<br>(n=112) | 0.89 |
| Six-minute walk distance, m | 375±125 (n=24) | 245±117 (n=65) | <0.0001 | 329±127 (n=97) | 218±100 (n=81) | <0.0001 |
| Echocardiography |  |  |  |  |  |  |
| RVFAC, % | 28±10 (n=34) | 27±10 (n=48) | 0.63 | 35±10 (n=92) | 32±11 (n=62) | 0.049 |
| TAPSE, cm | 1.8±0.5 (n=32) | 1.9±0.9 (n=54) | 0.93 | 1.9±0.6 (n=107) | 1.8±0.4 (n=75) | 0.47 |
| S', cm/s | 10.2±2.2 (n=22) | 10.6±3.7 (n=38) | 0.46 | 10.7±2.7 (n=78) | 11.2±2.4 (n=47) | 0.29 |
| Hemodynamics |  |  |  |  |  |  |
| Heart rate, beats/min | 74±15 (n=47) | 77±17 (n=87) | 0.36 | 79±15 (n=135) | 78±14 (n=95) | 0.60 |
| Mean right atrial pressure, mmHg | 7±4 (n=59) | 8±4 (n=99) | 0.20 | 8±5 (n=153) | 8±5 (n=111) | 0.90 |
| Mean pulmonary artery pressure, mmHg | 47±16 (n=60) | 40±10 (n=100) | 0.007 | 44±13 (n=156) | 42±12 (n=113) | 0.27 |
| Pulmonary capillary wedge pressure, mmHg | 10±3 (n=60) | 12±4 (n=98) | 0.007 | 9±4 (n=157) | 12±6 (n=111) | 0.001 |
| Cardiac output, L/min | 5.9±2.1 (n=43) | 5.1±1.5 (n=78) | 0.048 | 4.5±1.7 (n=112) | 4.8±1.4 (n=92) | 0.13 |
| Cardiac index, L/min/m<sup>2</sup> | 2.8±0.9 (n=41) | 2.5±0.9 (n=76) | 0.12 | 2.5±0.9 (n=112) | 2.6±0.7 (n=90) | 0.31 |
| Pulmonary vascular resistance, Wood units | 7.5±4.2 (n=53) | 6.5±3.4 (n=97) | 0.09 | 9.5±5.8 (n=150) | 7.2±3.8 (n=112) | 0.002 |
| Pulmonary arterial compliance, mL/mmHg | 2.2±1.5 (n=44) | 2.1±1.1 (n=86) | 0.85 | 1.6±1.0 (n=132) | 1.8±1.0 (n=95) | 0.17 |

COPD: chronic obstructive pulmonary disease, CPFE: combined pulmonary fibrosis and emphysema, CTD: connective tissue disease, HIV: human immunodeficiency virus, ILD: interstitial lung disease, NT-proBNP: N-terminal pro-brain natriuretic peptide, OSA: obstructive sleep apnea, PAH: pulmonary arterial hypertension, PH: pulmonary hypertension, PVOD: pulmonary veno-occlusive disease, RVFAC: right ventricular fractional area change, S': tricuspid annular peak systolic velocity, TAPSE: tricuspid annular plane systolic excursion.

Consistent with previous results(6), males had more impaired RV function than females in both PH types (**Figure 1A-B**). Next, we evaluated how PH etiology affected RV function in both sexes. There were no differences in RVFAC (Group 3: 27±10% vs. Group 1: 28±10%, *p*=0.63) or RVFAC relative to PVR (*p*=0.08 between y-intercepts, *p*=0.74 between slopes) between Group 3 PH and PAH males (**Figure 1C-D**). In contrast, female Group 3 PH patients had significantly lower RVFAC compared to female PAH patients (Group 3: 32±11% vs. Group 1: 35±10%, *p*=0.049) (**Figure 1E**). Group 3 females exhibited heightened sensitivity to RV afterload when compared to PAH females as RVFAC was lower at all PVR levels, and they had a more accentuated reduction in RV function as PVR increased (*p*=0.0005 between y-intercepts and *p*=0.05 between slopes) (**Figure 1F**). Importantly, postmenopausal (age≥55) Group 3 females had more severe RVD than postmenopausal PAH females when PVR was matched (**Figure 2**).

**Figure 1:**
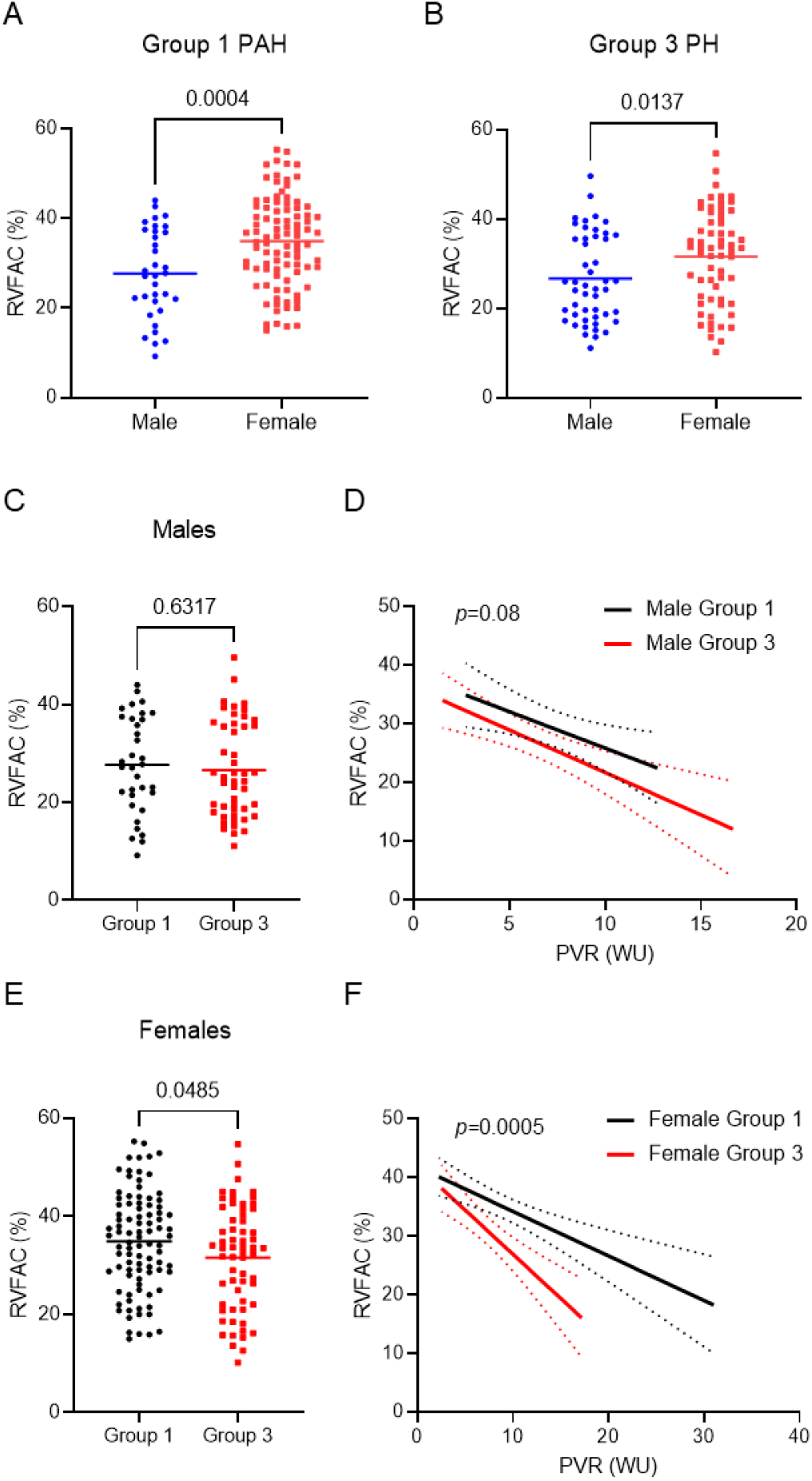
Group 3 females have exaggerated RV dysfunction compared to PAH females. Males with Group 1 PAH (**A**) and Group 3 PH (**B**) had significantly decreased RVFAC compared to females. (**C**) Group 3 PH males had similar RVFAC compared to Group 1 PAH males. (**D**) There was no statistical difference in the relationship between RVFAC and PVR between Group 1 and Group 3 males (*p*=0.08 when comparing y-intercepts and *p*=0.74 when comparing slopes). (**E**) Female Group 3 PH patients had significantly worse RV function compared to female Group 1 PAH patients. (**F**) Group 3 PH females had inferior RV function at all PVR levels (*p*=0.0005 when comparing y-intercepts and *p*=0.05 when comparing slopes). PAH: pulmonary arterial hypertension, PH: pulmonary hypertension, PVR: pulmonary vascular resistance, RVFAC: right ventricular fractional area change, WU: Wood units.

**Figure 2:**
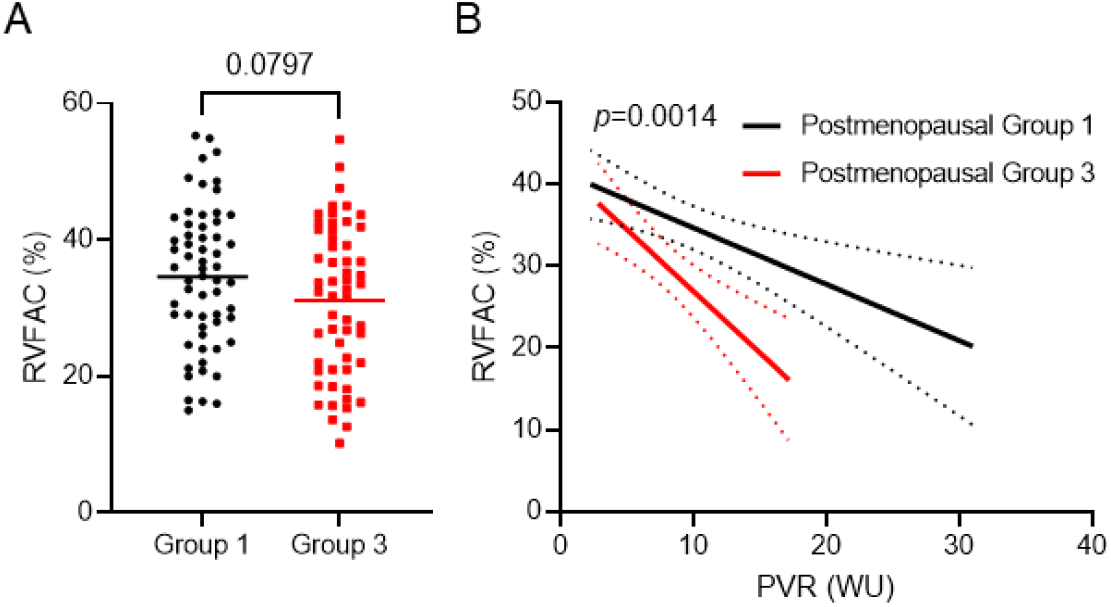
Postmenopausal Group 3 females have worse RV function compared to postmenopausal PAH females. (**A**) There was a non-statistically significant reduction in RVFAC between postmenopausal Group 3 females as compared to postmenopausal PAH females (Group 3: 31.1% [10.2-54.7%] vs. Group 1: 34.6% [15.0-55.3%], data presented as mean [range]). (**B**) Postmenopausal Group 3 PH females had inferior RV function at all PVR levels (*p*=0.0014 when comparing y-intercepts and *p*=0.06 when comparing slopes).

## Discussion

Here, we show females with Group 3 PH have exaggerated RVD when compared to PAH females, but male sex was not associated with differences in RV phenotype when comparing the two PH types. The biological basis for worse RV function in Group 3 PH females is uncertain, but age and the presence of lung disease are likely contributors. The older age in Group 3 females would lead to a higher proportion of postmenopausal patients, and that would ultimately result in lower estradiol levels. Estradiol is known to augment RV function as higher estradiol levels are associated with enhanced RV function(5). Moreover, preclinical studies show estrogen directly preserves RV function in rodent PAH(9, 10). However, potential differences in estradiol may not be the only contributor to worse RV function in Group 3 females as postmenopausal Group 3 females had more severe RVD compared to postmenopausal PAH patients. Thus, the presence of chronic lung disease likely exacerbates RVD in females with Group 3 PH.

Our study has important limitations that must be acknowledged. First, this was a single-center study. Second, we were unable to calculate RVFAC for all patients due to the quality of the echocardiographic images. Third, we may not have had the power to detect differences in RV function in males as there was a trend for worse RV function in male Group 3 PH compared to PAH males when we plotted RVFAC relative to PVR. Finally, our sample size was too small to determine whether there are differences in RV function between pre- and postmenopausal Group 3 females. We only had RVFAC data for six peri-to premenopausal (<50 years) Group 3 females, but there was similar RVFAC when compared to 32 PAH females <50 years (Group 3: 35.7±4.7% vs. Group 1: 35.5±9.7%).

In summary, we show female Group 3 PH patients have inferior RV function compared to female PAH patients. Chronic lung disease and decreased estradiol levels due to aging likely underlie these findings.

## Data Availability

All data are available from the authors upon request.

## Author Contributions

All authors made substantial contributions to the conception and design of the work. SZP, FK, and KWP completed the data acquisition and analysis. SZP and KWP drafted the manuscript. All authors revised the manuscript and approved the final version of the manuscript.

## List of non-standard abbreviations

mPAP: Mean pulmonary arterial pressure
PAC: Pulmonary arterial compliance
PAH: Pulmonary arterial hypertension
PH: Pulmonary hypertension
PVR: Pulmonary vascular resistance
RV: Right ventricle/ventricular
RVD: Right ventricular dysfunction
RVFAC: Right ventricular fractional area change

---

**Supplemental Table:**
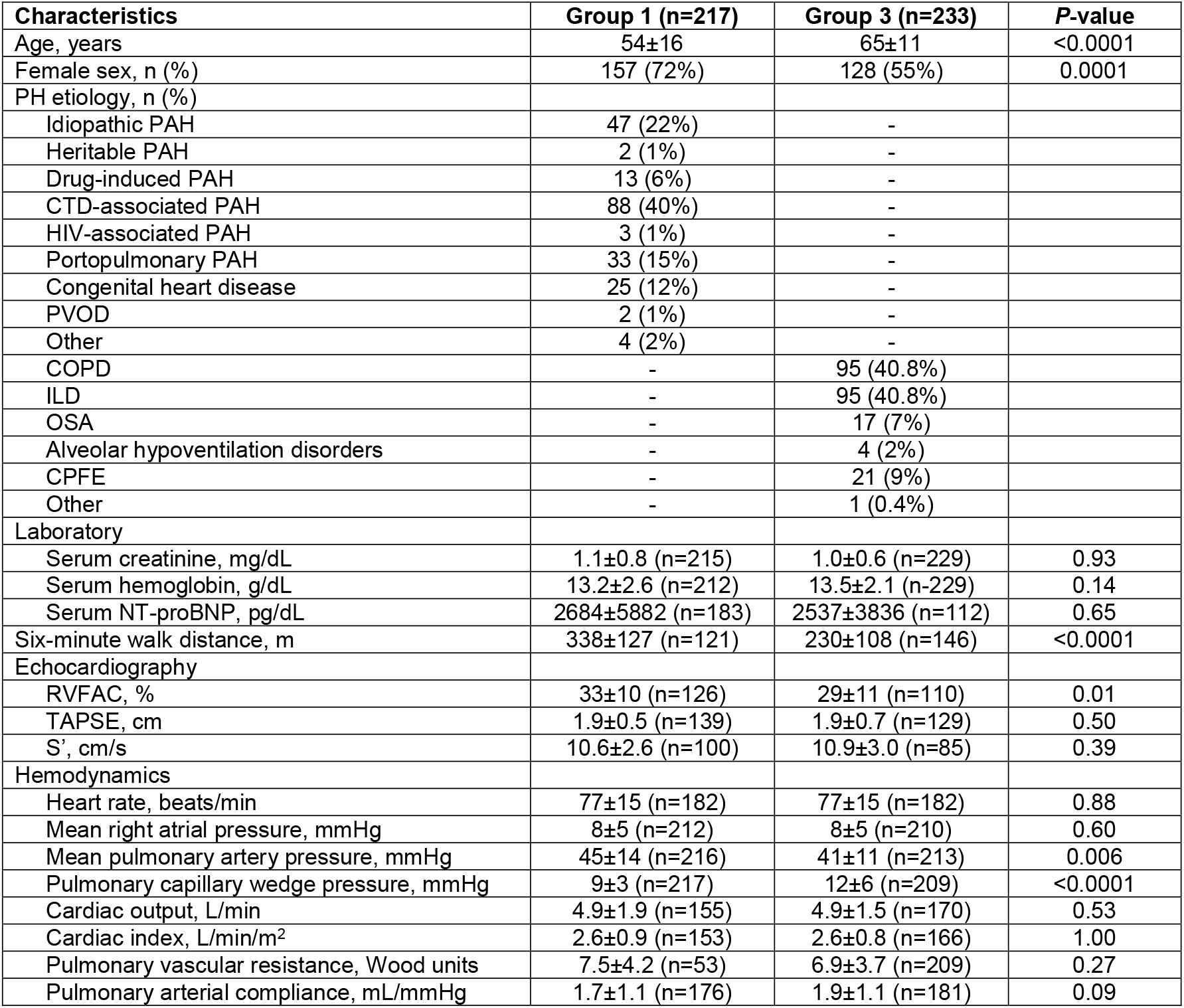
Clinical characteristics of the Group 1 PAH and Group 3 PH cohorts.

| Characteristics | Group 1 (n=217) | Group 3 (n=233) | P-value |
| --- | --- | --- | --- |
| Age, years | 54±16 | 65±11 | <0.0001 |
| Female sex, n (%) | 157 (72%) | 128 (55%) | 0.0001 |
| PH etiology, n (%) |  |  |  |
| Idiopathic PAH | 47 (22%) | - |  |
| Heritable PAH | 2 (1%) | - |  |
| Drug-induced PAH | 13 (6%) | - |  |
| CTD-associated PAH | 88 (40%) | - |  |
| HIV-associated PAH | 3 (1%) | - |  |
| Portopulmonary PAH | 33 (15%) | - |  |
| Congenital heart disease | 25 (12%) | - |  |
| PVOD | 2 (1%) | - |  |
| Other | 4 (2%) | - |  |
| COPD | - | 95 (40.8%) |  |
| ILD | - | 95 (40.8%) |  |
| OSA | - | 17 (7%) |  |
| Alveolar hypoventilation disorders | - | 4 (2%) |  |
| CPFE | - | 21 (9%) |  |
| Other | - | 1 (0.4%) |  |
| Laboratory |  |  |  |
| Serum creatinine, mg/dL | 1.1±0.8 (n=215) | 1.0±0.6 (n=229) | 0.93 |
| Serum hemoglobin, g/dL | 13.2±2.6 (n=212) | 13.5±2.1 (n=229) | 0.14 |
| Serum NT-proBNP, pg/dL | 2684±5882 (n=183) | 2537±3836 (n=112) | 0.65 |
| Six-minute walk distance, m | 338±127 (n=121) | 230±108 (n=146) | <0.0001 |
| Echocardiography |  |  |  |
| RVFAC, % | 33±10 (n=126) | 29±11 (n=110) | 0.01 |
| TAPSE, cm | 1.9±0.5 (n=139) | 1.9±0.7 (n=129) | 0.50 |
| S', cm/s | 10.6±2.6 (n=100) | 10.9±3.0 (n=85) | 0.39 |
| Hemodynamics |  |  |  |
| Heart rate, beats/min | 77±15 (n=182) | 77±15 (n=182) | 0.88 |
| Mean right atrial pressure, mmHg | 8±5 (n=212) | 8±5 (n=210) | 0.60 |
| Mean pulmonary artery pressure, mmHg | 45±14 (n=216) | 41±11 (n=213) | 0.006 |
| Pulmonary capillary wedge pressure, mmHg | 9±3 (n=217) | 12±6 (n=209) | <0.0001 |
| Cardiac output, L/min | 4.9±1.9 (n=155) | 4.9±1.5 (n=170) | 0.53 |
| Cardiac index, L/min/m<sup>2</sup> | 2.6±0.9 (n=153) | 2.6±0.8 (n=166) | 1.00 |
| Pulmonary vascular resistance, Wood units | 7.5±4.2 (n=53) | 6.9±3.7 (n=209) | 0.27 |
| Pulmonary arterial compliance, mL/mmHg | 1.7±1.1 (n=176) | 1.9±1.1 (n=181) | 0.09 |

